# Economic Benefits of COVID-19 Screening Tests

**DOI:** 10.1101/2020.10.22.20217984

**Authors:** Andrew Atkeson, Michael Droste, Michael J. Mina, James H. Stock

## Abstract

We assess the economic value of screening testing programs as a policy response to the ongoing COVID-19 pandemic. We find that the fiscal, macroeconomic, and health benefits of rapid SARS-CoV-2 screening testing programs far exceed their costs, with the ratio of economic benefits to costs typically in the range of 2-15 (depending on program details), not counting the monetized value of lives saved. Unless the screening test is highly specific, however, the signal value of the screening test alone is low, leading to concerns about adherence. Confirmatory testing increases the net economic benefits of screening tests by reducing the number of healthy workers in quarantine and by increasing adherence to quarantine measures. The analysis is undertaken using a behavioral SIR model for the United States with 5 age groups, 66 economic sectors, screening and diagnostic testing, and partial adherence to instructions to quarantine or to isolate.

## 1. Introduction

A now-large body of economic research concludes that much of the decline in economic activity associated with the SARS-CoV-2 pandemic is a consequence of self-protective behavior.^1^ An implication is that, for a robust pre-vaccine recovery, consumers must feel safe shopping, workers must feel safe returning to work, and parents must feel safe sending their children back to school. But to date, the public health policies that have been pursued in the United States – lockdowns, masks, social distancing, workplace safety, protecting the elderly, and so forth – have failed to suppress the virus, and the economic recovery has plateaued.

One yet-unused public health tool for suppressing the virus is widespread screening testing. Screening testing potentially can help control the virus by detecting and isolating contagious individuals who are asymptomatic, mildly symptomatic, or presymptomatic. Developing and deploying inexpensive rapid screening tests has been advocated since early in the pandemic, see for example Gottlieb et al. (2020), National Governors’ Association (2020), The Conference Board (2020), Romer (2020), Rockefeller Institute (2020), Silcox et al (2020), and Kotlikoff and Mina (2020). While the capacity for standard laboratory-based PCR tests for SARS-CoV-2 has increased, their cost, turnaround time, and the need to ensure PCR availability for diagnostic purposes constrain the ability of PCR testing to expand for widespread screening. Instead, new inexpensive rapid-turnaround tests have the potential for widespread use. These tests, however, exhibit lower sensitivity and specificity than laboratory PCR testing. In an editorial in the *Journal of Clinical Microbiology*, Pettengill and McAdam (2020) raise concerns that low specificity (that is, a high rate of false positives) would undercut the credibility of the screening program, reducing adherence to instructions to isolate if positive. Even with partial adherence to quarantine and isolation, low specificity would be a drag on the economy by placing many healthy workers in isolation. Pettengill and McAdam (2020) also point out that low sensitivity allows infected individuals to slip through the cracks. These concerns raise questions about the public health and economic benefits of imperfect screening tests.

This paper undertakes a macroeconomic cost-benefit analysis of a hypothetical U.S. program of imperfect screening testing with partial adherence. We extend the behavioral SIR model in Baqaee, Farhi, Mina, and Stock (2020b) (BFMS) to incorporate diagnostic and screening test regimes and partial adherence to self-isolation. Spread of the virus depends on contacts, which depend on economic activity; conversely, economic activity depends on the spread and trajectory of the virus. This model allows us to characterize how a given testing regime impacts the joint dynamics of disease transmission and economic activity.

The effectiveness of a screening testing program hinges on whether those who test positive adhere to the instruction to self-isolate. Using survey data from the United Kingdom covering March through August 2020, Smith et al. (2020) found that, of individuals reporting COVID symptoms, only 18% report self-isolating; among those who were told by the National Health Service that they had been in close contact with a confirmed COVID-19 case, only 11% reported quarantining for the recommended 14 days. These findings suggest that adherence will be low in response to other signals that have low information content – in particular, a low positive predictive value (PPV) ^2^ – about whether the individual is actually infected. We therefore allow the rate of adherence to depend on the specificity of the screening test.

Table 1 presents results for three representative testing programs. The programs are calibrated to existing or proposed tests and are designed to be representative of ones that might be deployable with adequate resources and effort. For cost-benefit purposes, we assume all incremental testing is federally funded.

**Table 1.**
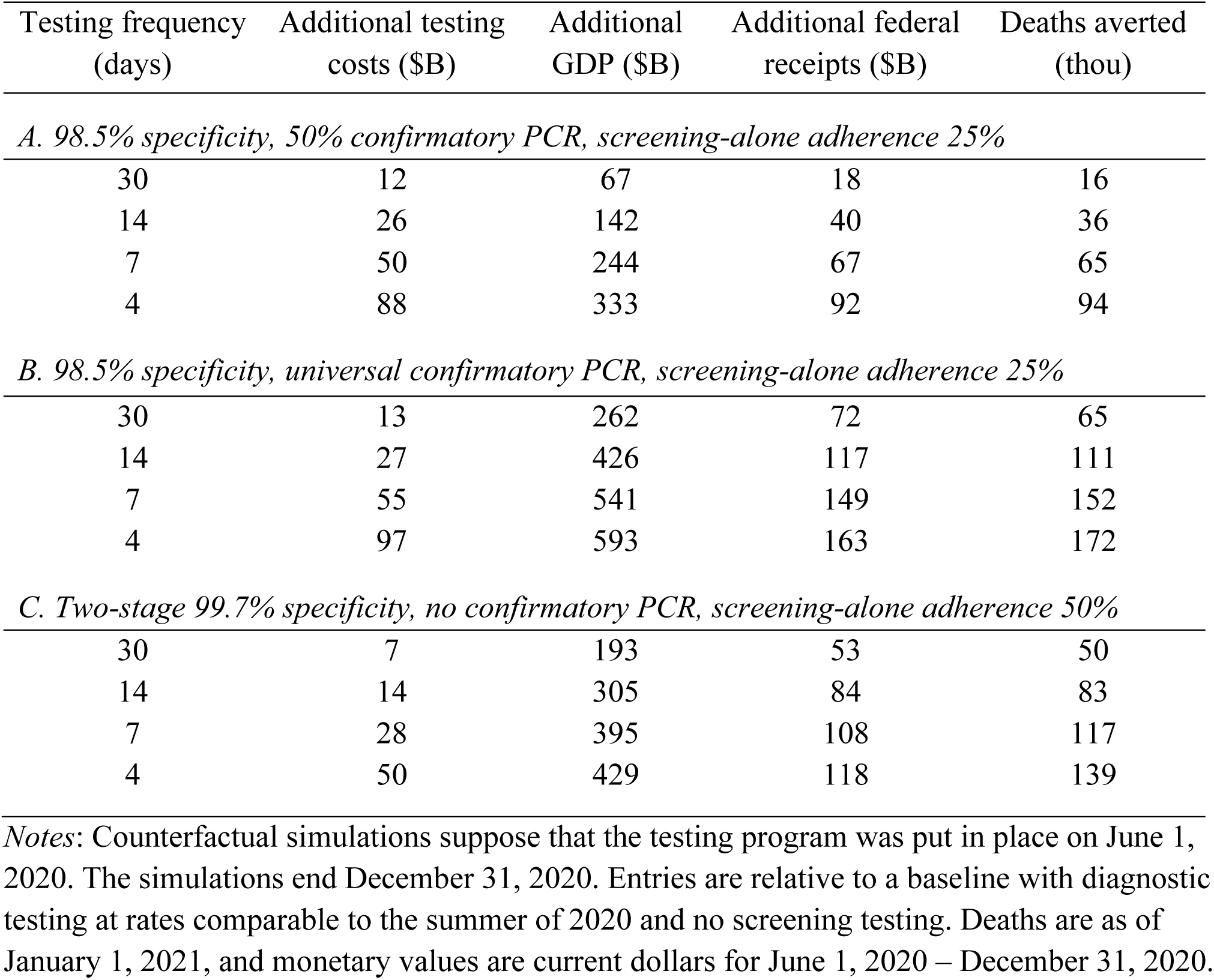
Economic and mortality impacts of three screening programs

Panel A considers a $5 screening test with 97.1% sensitivity and 98.5% specificity,^3^ in which half of those who test positive on the screening test take a $50 confirmatory PCR test with a 48-hour mean turnaround time. We suppose that adherence is high (75%) for those with a positive PCR test and low (25%) for those who test positive on the screening test but do not take a confirmatory PCR test. For random population screening testing at a weekly frequency, the total incremental cost of the program is $51 billion over the June 1 – December 31, 2020 simulation period. Using our epidemiological-economic model, we project 65,000 deaths averted, an increase in GDP of $244 billion, and an increase in federal tax revenues of $67 billion over the counterfactual period of the program, June 1 – December 31, 2020, relative to a baseline with diagnostic but not screening testing.

Panel B in Table 1 modifies this screening program so that everyone testing positive on the screening test receives a confirmatory PCR test. At a weekly testing cadence, this increases demand for PCR tests by approximately 630,000 tests per day, relative to the no-screening baseline. Testing costs are somewhat higher, but because the effective adherence rate is higher under this program than in Panel A, deaths averted rise to 152,000 and the increase in GDP is larger, $541 billion, for weekly testing.

Panel C considers a different screening testing program, with two-step testing with a combined two-step specificity of 99.7%. As an illustration, one way to achieve this specificity is by independent two-step rapid antigen tests as discussed by Mina, Parker, and Larremore (2020). The first step is a low-specificity (80%) $2 test, for example an inexpensive hypothetical paper-strip antigen test; if there is a positive test, the confirmatory test is the $5, 98.5% specificity test used in panels A and B.^4^ There is no confirmatory PCR testing. We suppose the 99.7% specificity evokes a 50% adherence rate. The incremental testing costs under this program are less than for programs A or B. In part because turnaround is rapid, it averts 117,000 deaths and increases GDP by $395 billion at a weekly testing frequency, despite assumed lower adherence than to a PCR-based regime.

These results and the additional sensitivity analysis below lead to four main conclusions.

First, even with partial compliance, screening testing induces large net economic benefits. For the cases in Table 1, economic benefits exceed costs by a factor of 5-10 for weekly testing. If all the tests were paid for by the federal government, the additional tax revenues generated by the induced GDP growth would more than pay for the testing costs. Net benefits rise if one additionally monetizes deaths averted using a statistical value of life.

Second, the signal value of a single positive screening test is low: in our simulations, the PPV is typically less than 5% for a test with 98.5% specificity. This low signal value could lead to low adherence and, among those who do adhere, imposes economic costs because of healthy workers isolating. Introducing confirmatory testing into the program increases the signal value, reduces unnecessary isolation, and arguably would lead to greater adherence, increasing net benefits.

Third, screening test sensitivity is of secondary importance – a finding that is consistent with, for example, Larremore et al (2020) and Paltiel, Zheng, and Walensky (2020). For example, the results in Table 1 are very similar if screening test sensitivity is reduced from 97.1% to 85%: even at 85% sensitivity, the vast majority of the tested infected are detected and, if they adhere, isolated.

Fourth, we find that targeting testing to younger and middle-aged adults can improve both economic and mortality outcomes, holding constant the number of screening tests. Although targeting those ages retards activity by sending workers into isolation, it breaks the chain of transmission to the elderly.

The model is summarized in Section 2. Section 3 presents the results for uniform testing, and age-based testing is examined in Section 4.

### Related literature

This paper is related to a growing literature synthesizing epidemiological models of disease transmission with macroeconomic dynamics,^5^ some of which considers testing and quarantine. Berger, Herkenhoff, and Mongey (2020) consider the effects of testing and quarantine in a SIR model with a single perfect test and imperfect adherence to quarantine; the authors show that testing can reduce the severity of lockdowns required to achieve a given reduction in the spread of disease. Cherif and Hasanov (2020) study the costs and returns of a test-and-quarantine strategy in a SIR model, paying particular attention to ‘smart’ testing strategies that take advantage of spatial heterogeneity in disease prevalence and population density. Brotherhood, Kircher, Santos, and Tertilt (2020) and Eichenbaum, Rebelo, and Trabandt (2020b) consider age-varying diagnostic testing and quarantine; in both models, the role of testing is primarily to resolve individual uncertainty about infection status. Other papers that address testing, contact tracing, and/or quarantine are Acemoglu, Makhdoumi, Malekian, and Ozdaglar (2020), Augenblick, Obermeyer, Kolstad, and Wang (2020), BMFS (2020b), Gans (2020), and Piguillem and Shi (2020). A closely related paper in the epidemiological literature is Paltiel, Zheng and Walensky (2020), who consider college coronavirus testing and incorporate costs of tests and of housing the quarantined. Also see, among others, Larremore et al (2020), Taiaple, Romer, and Linnarsson (2020) and Peto et al (2020).

Relative to this literature, our main contribution is to provide carefully calibrated and estimated model for assessing the net economic, fiscal, and total (including mortality) benefits of multi-step imperfect screening testing in conjunction with diagnostic testing. By combining a 66-sector economic model with a five-age behavioral SIR model, we can consider age-based strategies and the effect of temporary isolation on employment and output.

## 2. Behavioral SIR model with screening and diagnostic testing

Our starting point is the BFMS behavioral SIR model, which connects a SIR model of disease transmission to economic activity. The BFMS model has five age groups (ages 0-19, 20-44, 45-64, 65-74, and 75+) and 66 private economic sectors plus the government. Pre-pandemic contact matrices are estimated from POLYMOD (Mossong et. al. (2017)) for work, home, and other activities. Work contact matrices vary by sector depending on sectoral worker proximity (Mongey, Pilossoph and Weinberg (2020)). The behavioral aspect of the model arises from a feedback rule in which activity depends on the current weekly death rate and the slope of the weekly death rate. In addition, the behavioral rule has a lockdown-fatigue component in which high unemployment rates and cumulative past unemployment rates contribute (all else equal) to a desire to resume activity. Epidemiological parameters, including age-based death rates, are taken from the epidemiological literature, from the CDC, or, for the transmission rate and initial infection rate, estimated from US data on daily deaths. For additional details, see Appendix 1 and BFMS.

### 2.1 Extension to screening and diagnostic testing

This paper extends the BFMS model to incorporate explicit screening and diagnostic testing with partial adherence. The key elements of this extension are:

1. Individuals are selected at random, at a daily rate *μ*, for rapid screening testing.
2. A fraction *ν* of individuals testing positive in the screening test take a confirmatory diagnostic (PCR) test; the remaining fraction 1-*ν* of those who test positive are instructed to self-isolate.
3. Symptomatic individuals can receive a diagnostic test.
4. Those awaiting diagnostic test results are instructed to quarantine.
5. The isolation pool consists of those with a “terminal” positive test result: a positive test among the PCR-tested symptomatic, a positive PCR test among the fraction *ν* of screening-test positives who take a confirmatory test, or a positive screening test among the fraction 1-*ν* who do not.
6. Adherence to instructions to quarantine or to isolate is partial.

The extended SIR model is illustrated in Figure 1 (equations are given in Appendix 1). The horizontal flows represent the disease progression from susceptible to exposed to infected to recently recovered to fully recovered, or from infected to deceased. The distinction between recently recovered and fully recovered is that the recently recovered test positive on a PCR test but are not contagious, e.g. see Larremore et al. (2020). The screening test is assumed to be less sensitive than the PCR test so detects the virus among the infected but not among the recently recovered.

**Figure 1.**
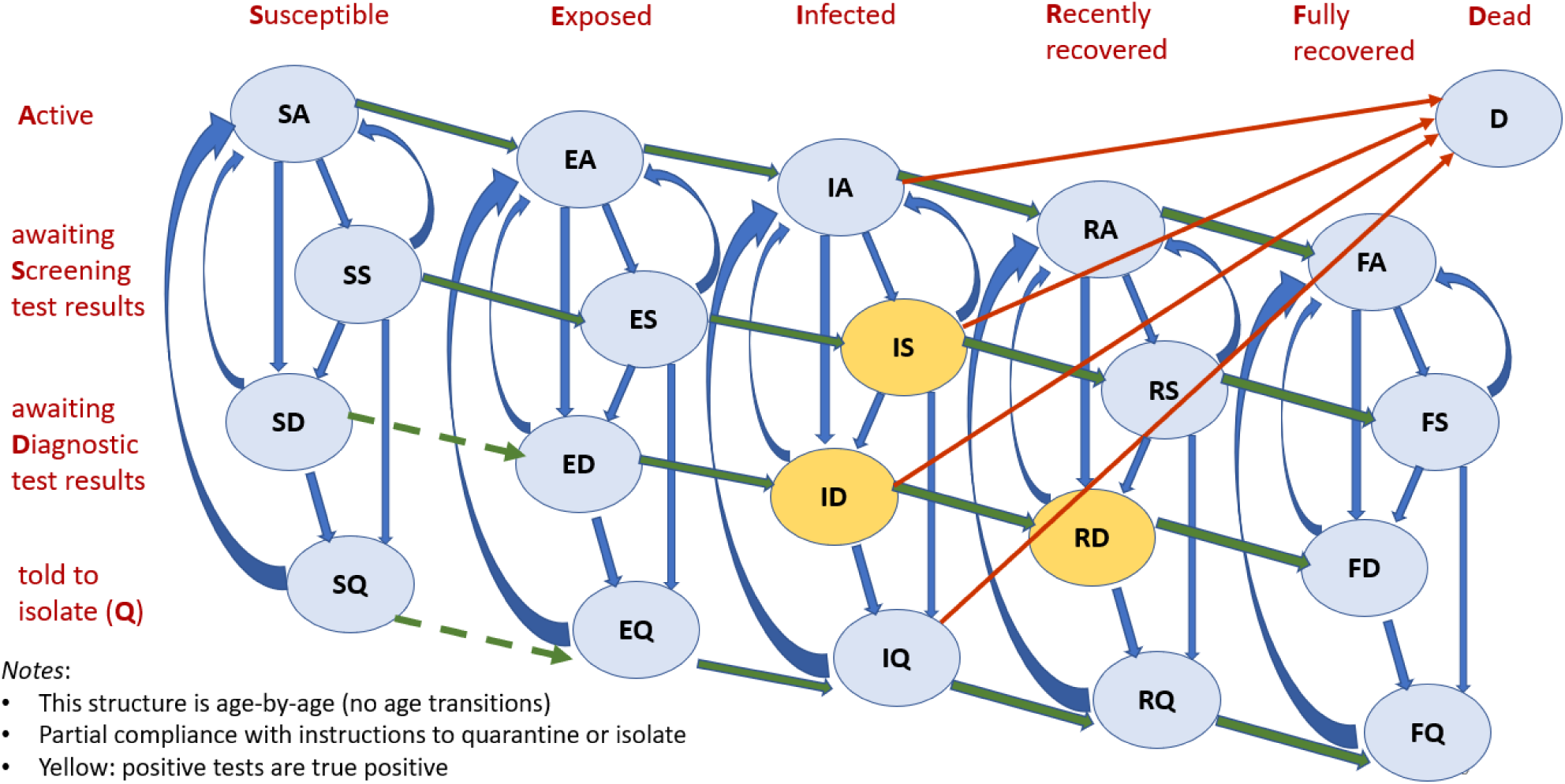
SIR model with screening & diagnostic testing and partial adherence.

Those instructed to isolate enter the isolation compartment (*Q*, to distinguish it from the infected *I*). The extent to which they adhere with that instruction depends on whether they arrived by testing positive on the diagnostic test, which has high signal value, or on the screening test, which has lower signal value. (Although we describe this as, say 25% of those arriving from a positive screening test as adhering, because of the homogenous structure of the model this is equivalent to all those arriving by this channel reducing their contacts by 25%.) With full adherence, the susceptible in quarantine and in isolation would not become infected, however because adherence is partial some of the isolated susceptibles (*SQ*) and quarantined susceptibles (*SD*) can become exposed.

Table 2 describes our baseline parameter values. The sensitivity and specificity of diagnostic test are intended to be in the range of laboratory PCR tests. The sensitivity and specificity of the baseline screening test are calibrated to analytical estimates corresponding to the BinaxNOW™ rapid antigen test, although we consider alternative values. The rate of uptake of diagnostic testing among the non-infected (*ρ*_0_) and among the infected (*ρ*_1_) were calibrated to match the total number of tests and the positivity rate in the US during July and August 2020.

**Table 2.** Testing Parameter Values

| Parameter | Definition | Value |
| --- | --- | --- |
| $p_S$ | Screening test: sensitivity | 0.97 |
| $q_S$ | Screening test: specificity | 0.985 |
| $p_D$ | Diagnostic test: sensitivity | 0.999 |
| $q_D$ | Diagnostic test: specificity | 0.997 |
| $\alpha_S$ | Isolation non-adherence rate after positive screening test | 0.75 for $q_S = 0.985$<br>0.5 for $q_S = 0.997$ |
| $\alpha_D$ | Isolation non-adherence rate after positive diagnostic test | 0.25 |
| $v$ | Fraction of screening positives who take confirmatory diagnostic test | 0, 0.5, 1 |
| $\lambda_D$ | Diagnostic test return rate ($1/\lambda_D$ is mean delay in days) | 1/2 |
| $\lambda_S$ | Screening test return rate | 1/.1 |
| $\mu$ | Screening testing frequency | 0,...,1/3 |
| $\rho_0$ | Rate that non-infected take diagnostic test | 1/600 |
| $\rho_1$ | Rate that infected take diagnostic test | 1/10 |
| $\zeta$ | Flow rate out of isolation ($Q$) | 1/14 |
| $\theta$ | Flow rate from recently to fully recovered | 1/5 |
| | Screening test cost | \$5, \$3 |
| | Diagnostic test cost | \$50 |
| | Value of a statistical life (USEPA 2020 in \$2020) | \$9.3m |

Notes: Alternative parameter values are considered as sensitivity analyses.

We allow the frequency of screening tests (governed by *μ*) to vary between 0 (no screening tests at all) to 0.3 (each person is screened on average every three days). Our baseline isolation adherence rate is 25% for those testing positive on the screening test with 98.5% specificity, is 50% for those testing positive on the two-stage screening test with 99.7% specificity, and is 75% for those testing positive on the diagnostic test (either after qualifying by being symptomatic or as the second stage following a positive screening test).

### 2.2 Simulation baseline and testing counterfactuals

BFMS has two baseline scenarios, both exhibiting a second wave of infections starting mid-summer 2020. In BFMS, the second wave was induced by a relaxation of social distancing, masks, and other protections, combined with a full return to school in the fall. The difference between the two scenarios was the strength of the feedback from deaths and the growth rate of deaths to activity. The baseline here uses feedback parameters that are a mid-point between the two baseline scenarios considered in BFMS.

We estimate the model using data through June 12, 2020. The simulation period begins June 1, 2020 and ends on January 1, 2021. Thus, the simulations reflect alternative, counterfactual paths for the virus and the economy for the final seven months of 2020.

Figure 2 shows the time path of actual deaths (black dashed), simulated deaths (red), and the level of GDP (green) indexed to its level in February 2020, under our baseline calibration with no screening testing. The bands for deaths and GDP are standard error bands based on estimation uncertainty for the model parameters. Although the baseline scenario was constructed in June, it closely tracks the path of deaths through mid-August. Subsequently, simulated deaths exceed actual deaths, in part because the simulation presumes a full return to school whereas many school districts chose remote or hybrid reopenings. Under the baseline, there are 359,000 deaths by January 1.

**Figure 2.**
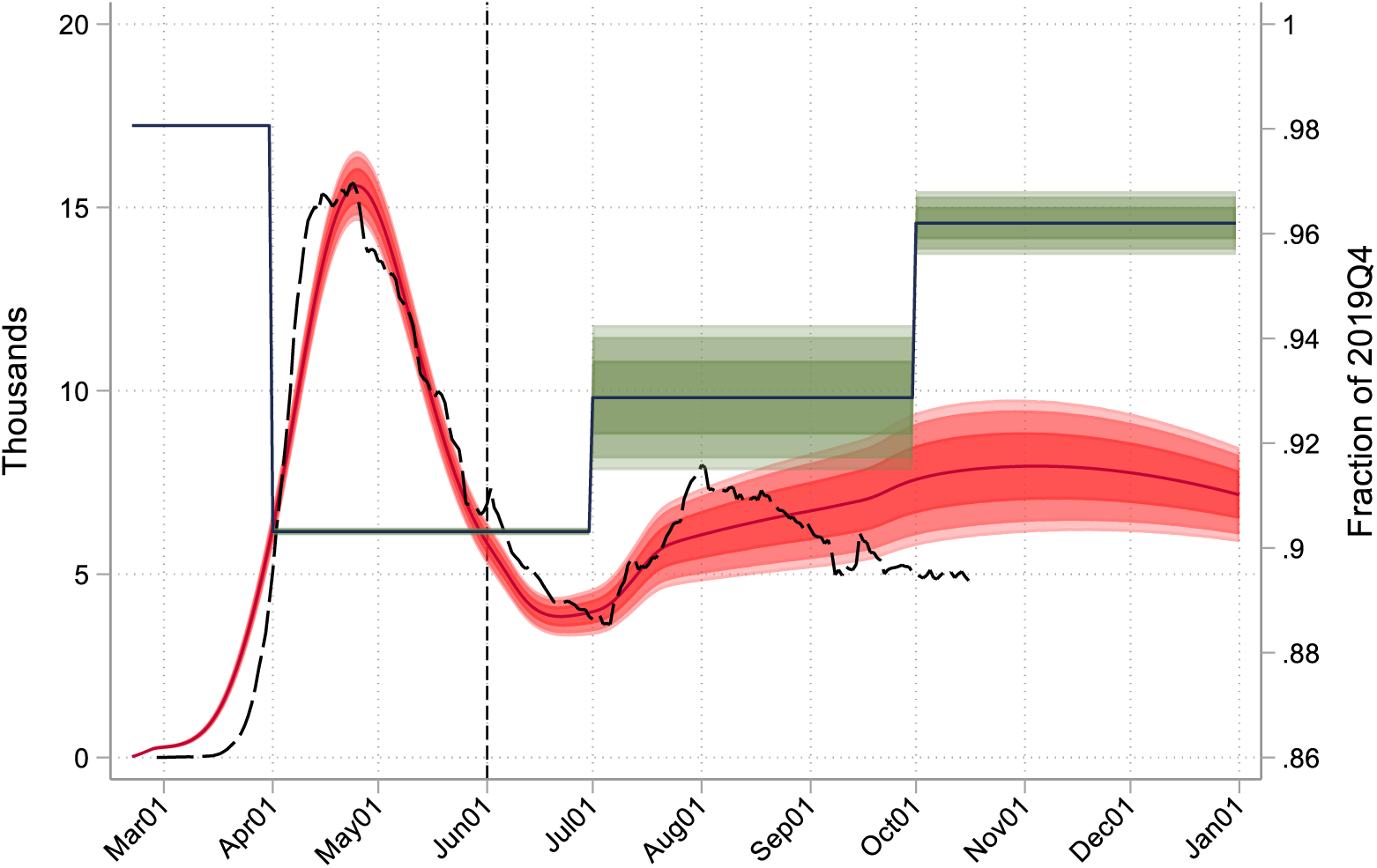
Actual and simulated paths for deaths and GDP: No-screening baseline. *Notes*: Quarterly GDP (green step function) is shown in real levels, indexed to 1 in 2019Q4. Total deaths (actual in black dashed, simulated in red) under the baseline simulation are 359,000 by January 1, 2021. Bands denote 67%, 90%, and 95% confidence bands using standard errors for the estimated model parameters.

Under the baseline, there are no screening tests (*μ* = 0), however there are diagnostic tests at rates that match the volume and positivity rates of actual testing in July and August. Under the screening testing counterfactuals, diagnostic testing is augmented by screening testing, holding constant all model parameters except for those describing the screening tests.

### 2.3 Evaluation of costs and benefits

The incremental costs and benefits of testing are computed from the number of tests and the model-implied economic and mortality outcomes under testing and no-testing scenarios.

We assume the cost of the 98.5% specific screening test is $5. The 80% specific screening test is assumed to cost $2, but is packaged for use with the $5 test with 98.5% specificity at a 5-to-1 ratio for an average cost of $3/test. The price of a diagnostic PCR test varies considerably in the United States; we use $50 for our baseline.

We compute three measures of benefits of tests: incremental GDP, incremental federal government revenues, and the monetized value of deaths avoided.

GDP is measured in 2020 dollars. Because the simulations start on June 1, GDP is the same under baseline and testing alternative have the same values for GDP for the first five months of the year, so any differences in GDP under the two scenarios occurs only from June through December; these incremental dollars of GDP are dollars for those seven months only, not at an annual rate.

The effect of an increase in GDP on government revenues is computed using elasticities of income taxes, corporate profits taxes, FICA, and the self-employment contributions tax from the Congressional Budget Office (Russek and Kowalewski (2015, Table 3) and CBO (2019)). Like GDP, these incremental revenues are for June-December only and are not annualized. To compute net fiscal benefits, we assume that all incremental testing is paid for by the Federal government.

**Table 3.** Economic and mortality impacts: Sensitivity analysis

| Testing frequency<br>(days) | Additional testing<br>costs (\$B) | Additional<br>GDP (\$B) | Additional federal<br>receipts (\$B) | Deaths averted<br>(thou) |
| --- | --- | --- | --- | --- |
| *D. Program A, except no confirmatory testing* |  |  |  |  |
| 30 | 11 | -10 | -3 | -2 |
| 14 | 23 | 56 | 15 | 17 |
| 7 | 46 | 166 | 46 | 48 |
| 4 | 79 | 266 | 73 | 84 |
| *E. Program A, except 97% screening test specificity* |  |  |  |  |
| 30 | 13 | 4 | 1 | 2 |
| 14 | 27 | 83 | 23 | 24 |
| 7 | 53 | 201 | 56 | 58 |
| 4 | 91 | 294 | 81 | 93 |
| *F. Program A, except screening-alone adherence 50%* |  |  |  |  |
| 30 | 12 | 171 | 47 | 45 |
| 14 | 25 | 288 | 78 | 82 |
| 7 | 51 | 365 | 100 | 120 |
| 4 | 88 | 363 | 100 | 142 |
| *G. Program B, except \$100 diagnostic test cost* | | | | |
| 30 | 14 | 262 | 72 | 65 |
| 14 | 31 | 429 | 117 | 111 |
| 7 | 63 | 544 | 149 | 152 |
| 4 | 109 | 593 | 164 | 172 |

Notes: See the notes to Table 1.

Deaths avoided are the cumulative number of deaths from COVID-19 on January 1, 2020 under the testing scenario, minus the total number of deaths in the baseline scenario. Deaths are monetized using the value of statistical life is from the US EPA (2020), converted to 2020 dollars, which is $9.3 million per life.

## 3. Results

This section provides full results for the three programs in Table 1, then provides sensitivity checks and time paths of the virus and GDP for illustrative programs.

### 3.1 Single-stage screening with partial confirmatory testing

We begin with program A in Table 1, a single-stage screening test with 98.5% specificity with 50% confirmatory PCR testing. The adherence rates are 25% for those instructed to isolate based on the screening test alone and 75% for those instructed to isolate based on the diagnostic test.

Mortality and economic outcomes for program A in Table 1 are shown in Figure 3. In all figures, the outcome of interest is plotted as a function of the screening test intensity *μ*. The multiple lines in the figures represent different screening test sensitivities, from 80% to 98.5%. Relative to the no screening test baseline, testing biweekly is estimated to avert approximately 37,000 deaths, and testing weekly averts 66,000 deaths, when screening test sensitivity is 97%. The number of days that individuals are told to isolate (upper right) increases approximately linearly with the amount of screening testing (there is some curvature because symptomatic testing falls as screening intensity increases). For weekly testing, there are approximately 930 million proscribed isolation days, which amounts to 1.3% of the total of 70 billion person-days during the June-December simulation period. (Because of partial adherence, only some of those isolation-days are actually observed.) Because only half of the screening-testing positives receive confirmatory PCR testing, the preponderance of those instructed to isolate are false positives. The screening test PPV (middle left) is low, approximately 6% for weekly testing. Because the virus is increasingly suppressed as *μ* increases, GDP (middle right) increase with *μ* for low and moderates testing rates *μ*; although the increase is held back by the large number of healthy workers who are isolating.

**Figure 3.**
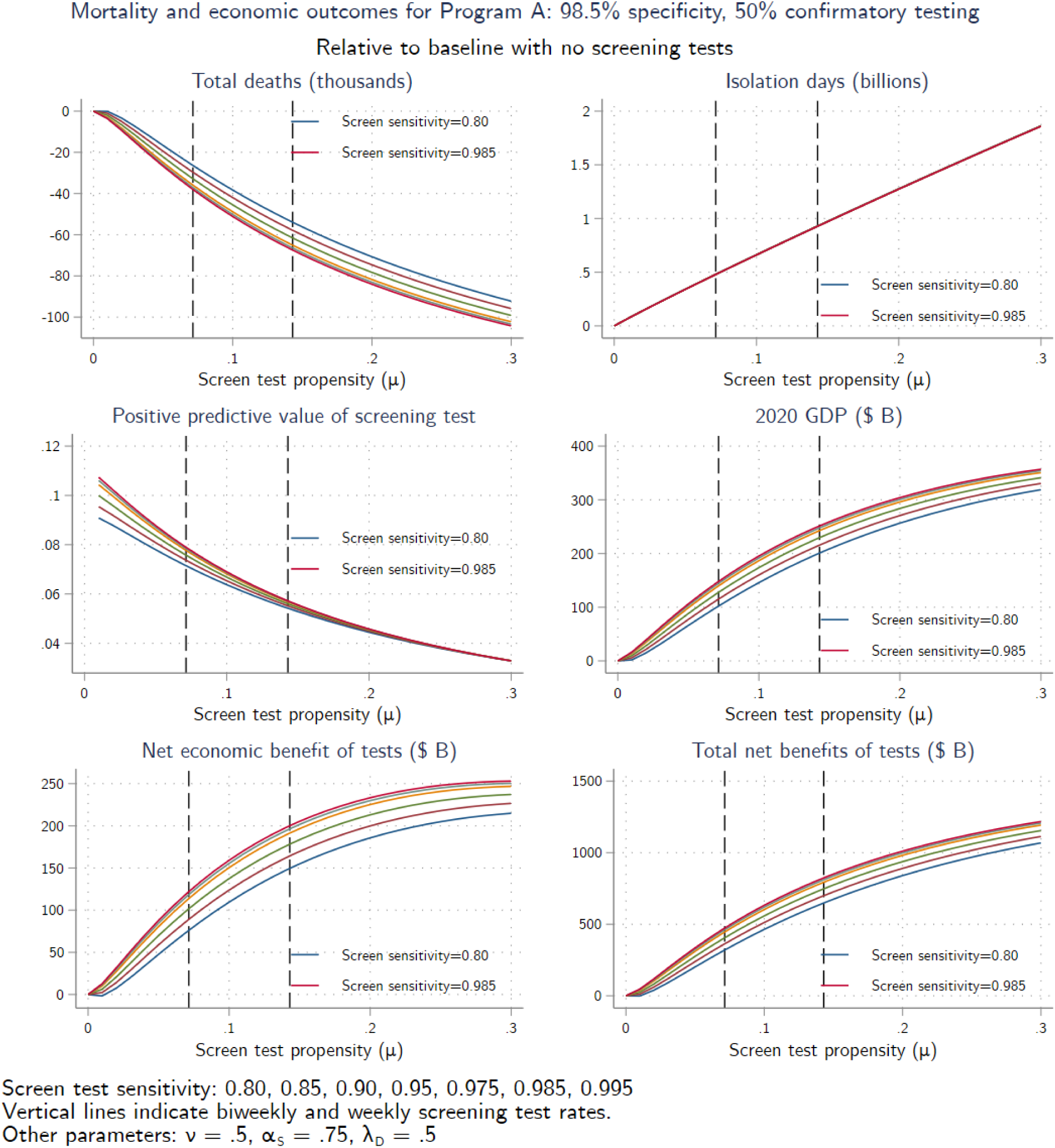
Mortality and economic outcomes for program A (50% confirmatory testing)

Cost-benefit results screening program A are shown in the bottom panel of Figure 3 and in Figure 4. Additional testing costs rise approximately linearly with the testing rate. For these parameter values, net economic benefits (Figure 3, lower left) are in the range of $75-120 billion for biweekly testing and $150-200 billion for weekly testing, depending on the screening test sensitivity. When the value of life is included as a benefit (Figure 3, lower right), net benefits are in the range of $320-470 billion for biweekly testing and $650-820 billion 215 for weekly testing, depending on the screening test sensitivity. In nearly every case considered, the screening program pays for itself (Figure 4, middle right), under the assumption that all additional testing is paid for by the federal government.

**Figure 4.**
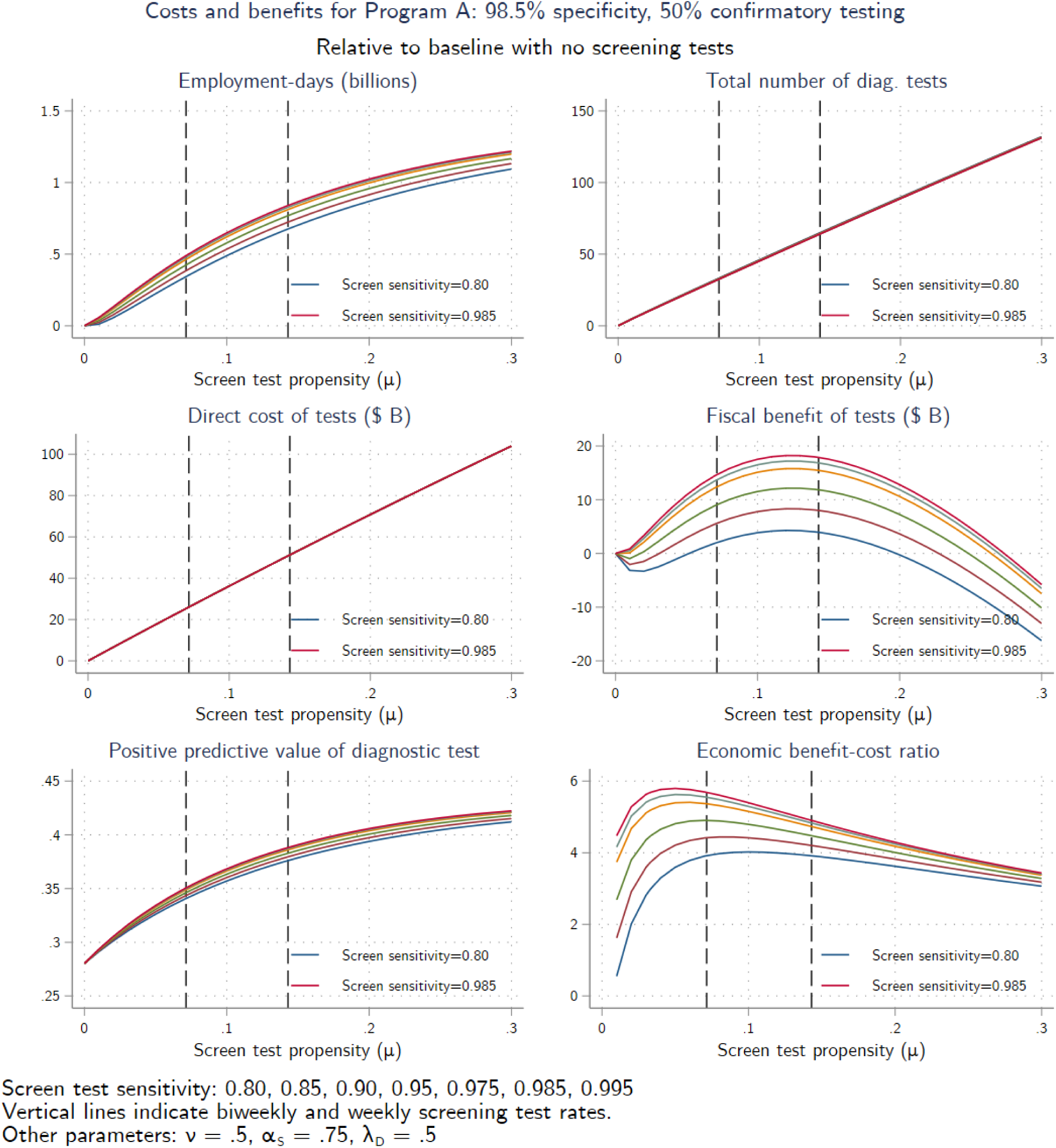
Costs and benefits for program A (50% confirmatory PCR testing)

In all these figures, increasing the sensitivity of the screening test improves outcomes, but those improvements are typically small relative to the gains from introducing the screening program in the first place.

### 3.2 Universal access to confirmatory PCR testing

Figure 5 and Figure 6 are the counterparts of Figure 3 and Figure 4 for program B, which has the same screening test but with universal confirmatory PCR testing. Expanding confirmatory testing from 50% to 100% substantially reduces deaths and increases GDP, because of the assumed greater adherence rate for highly specific PCR testing than for the screening test alone. In addition, because universal confirmatory testing reduces the number of healthy individuals in isolation; avoiding isolating the healthy allows them to work, increasing GDP. In fact, despite the increase in testing, isolation days are less under program B than under the no-screening baseline because the prevalence of the virus is substantially reduced, reducing the total number of positive diagnostic tests despite the inflow from positive screening tests. Universal confirmatory testing increases testing costs, so it is not obvious *a-priori* whether offering universal confirmatory testing increases or decreases net economic benefits; for the values considered here, the economic benefits of universal testing dominate and net economic benefits increase.

**Figure 5.**
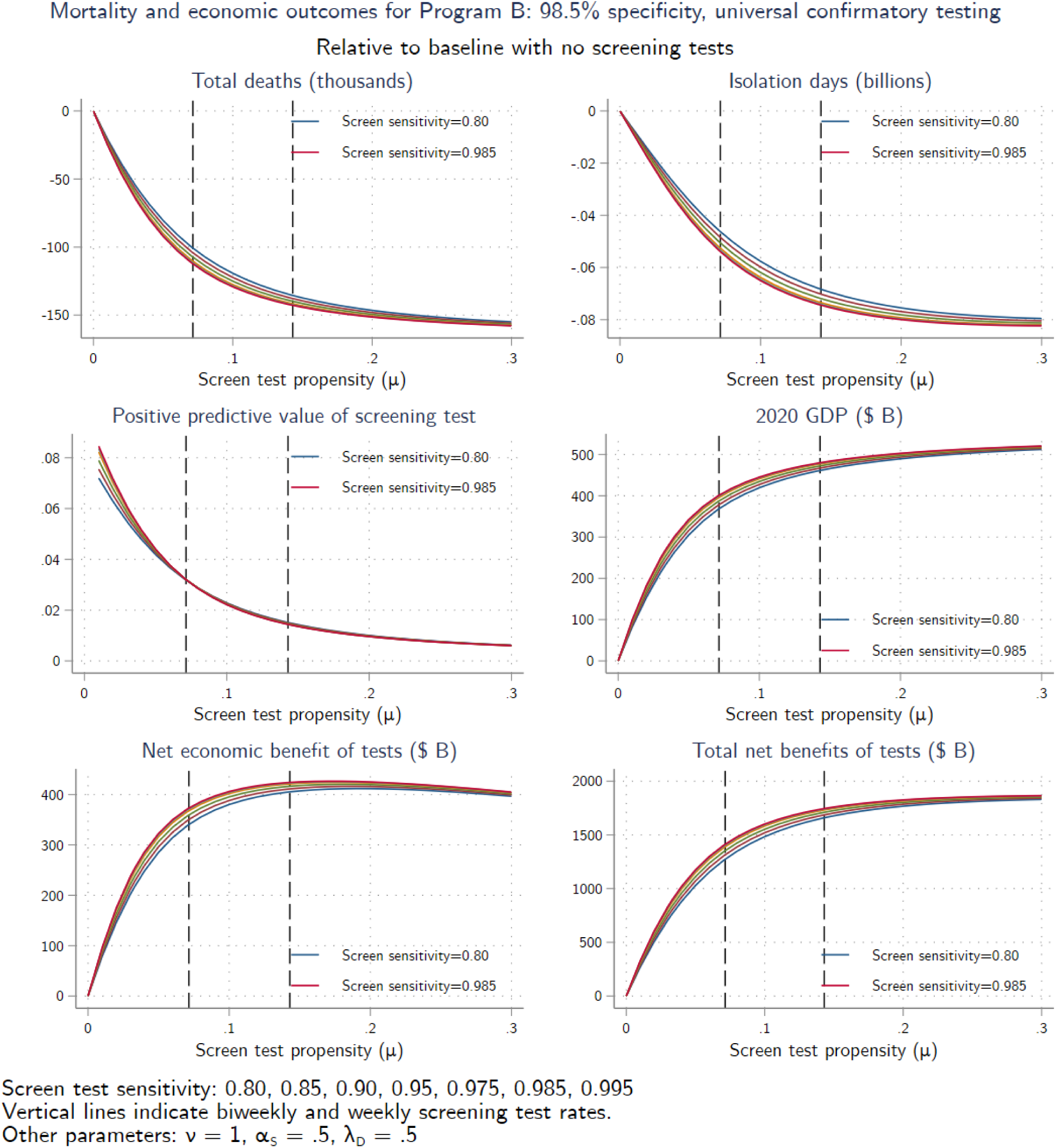
Mortality and economic outcomes for program B (universal confirmatory testing)

**Figure 6.**
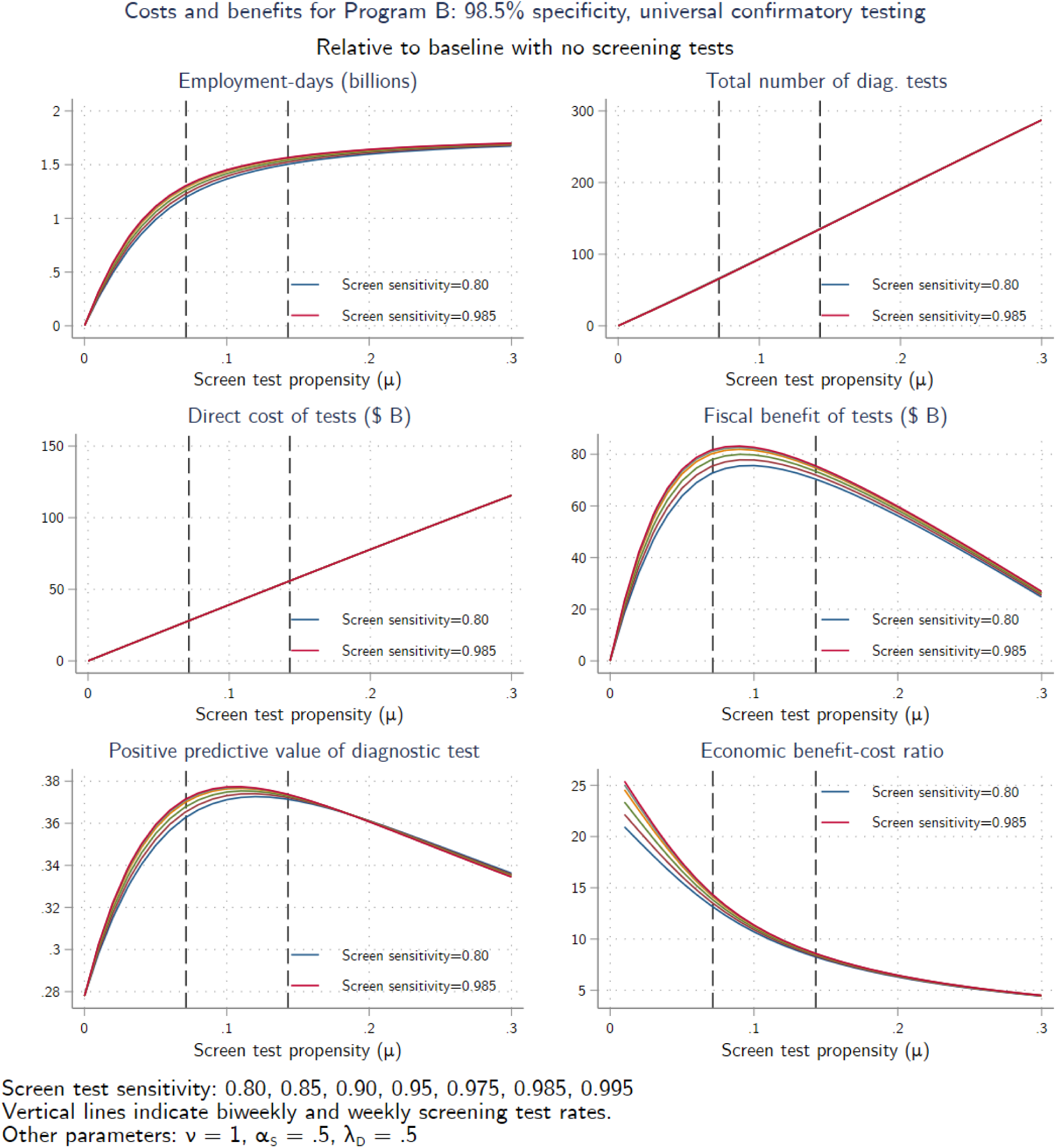
Costs and benefits for program B (universal confirmatory PCR testing)

### 3.3 Two-step screening with 99.7% specificity

Results for the two-step screening test of program C are shown in Figure 7 and Figure 8. This program has a $3 two-step screening test with specificity of 98.5%, adherence of 50%, and no confirmatory PCR testing. Mortality gains, proscribed isolation days, employment gains, and GDP gains fall between the tests in programs A and B, a consequence of the assumed lower adherence rates. Although net economic benefits are less for program C than for program B, the benefit-cost ratios are greatest for program C because the tests are less expensive.

**Figure 7.**
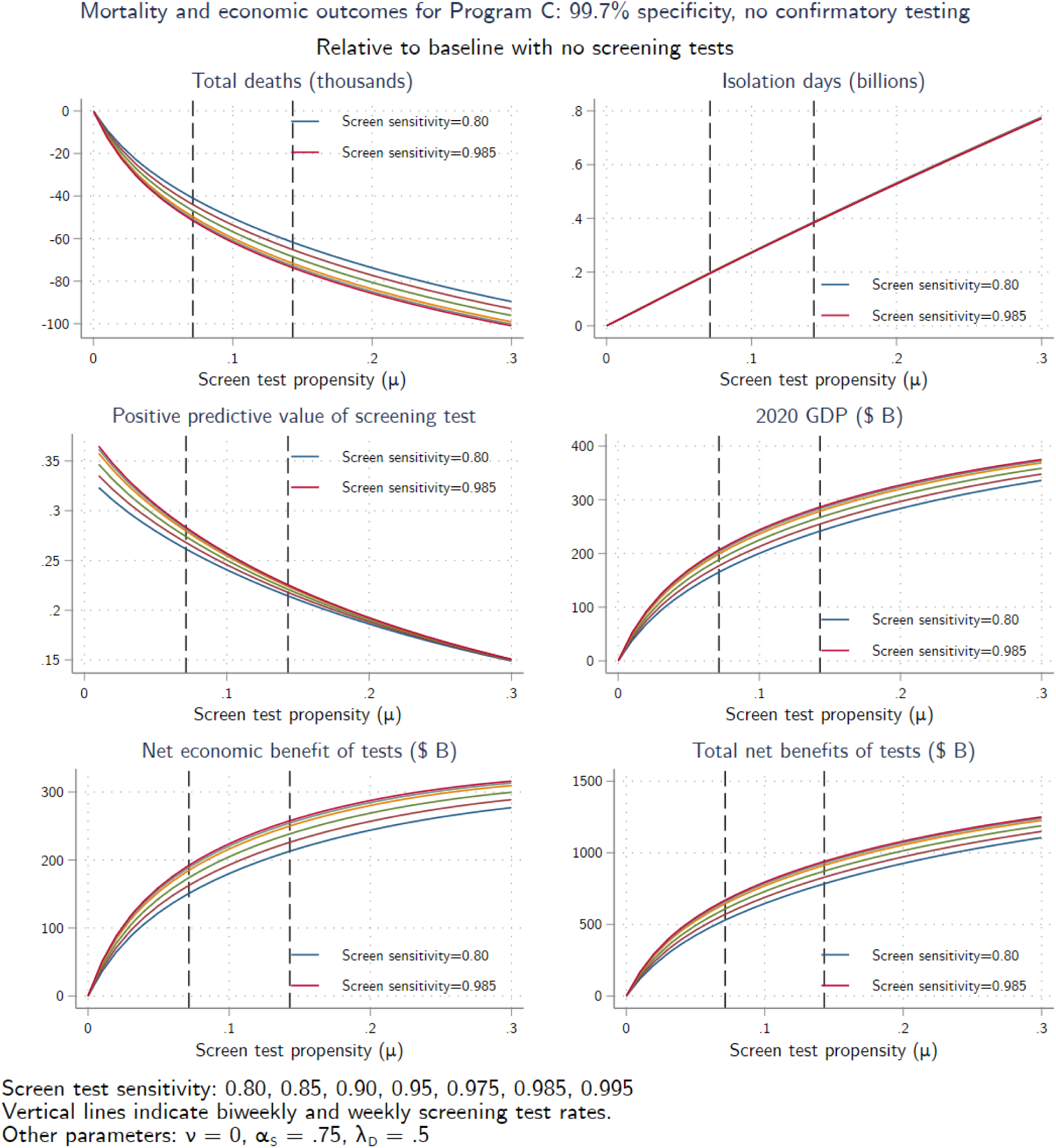
Mortality and economic outcomes for program C (no confirmatory PCR testing)

**Figure 8.**
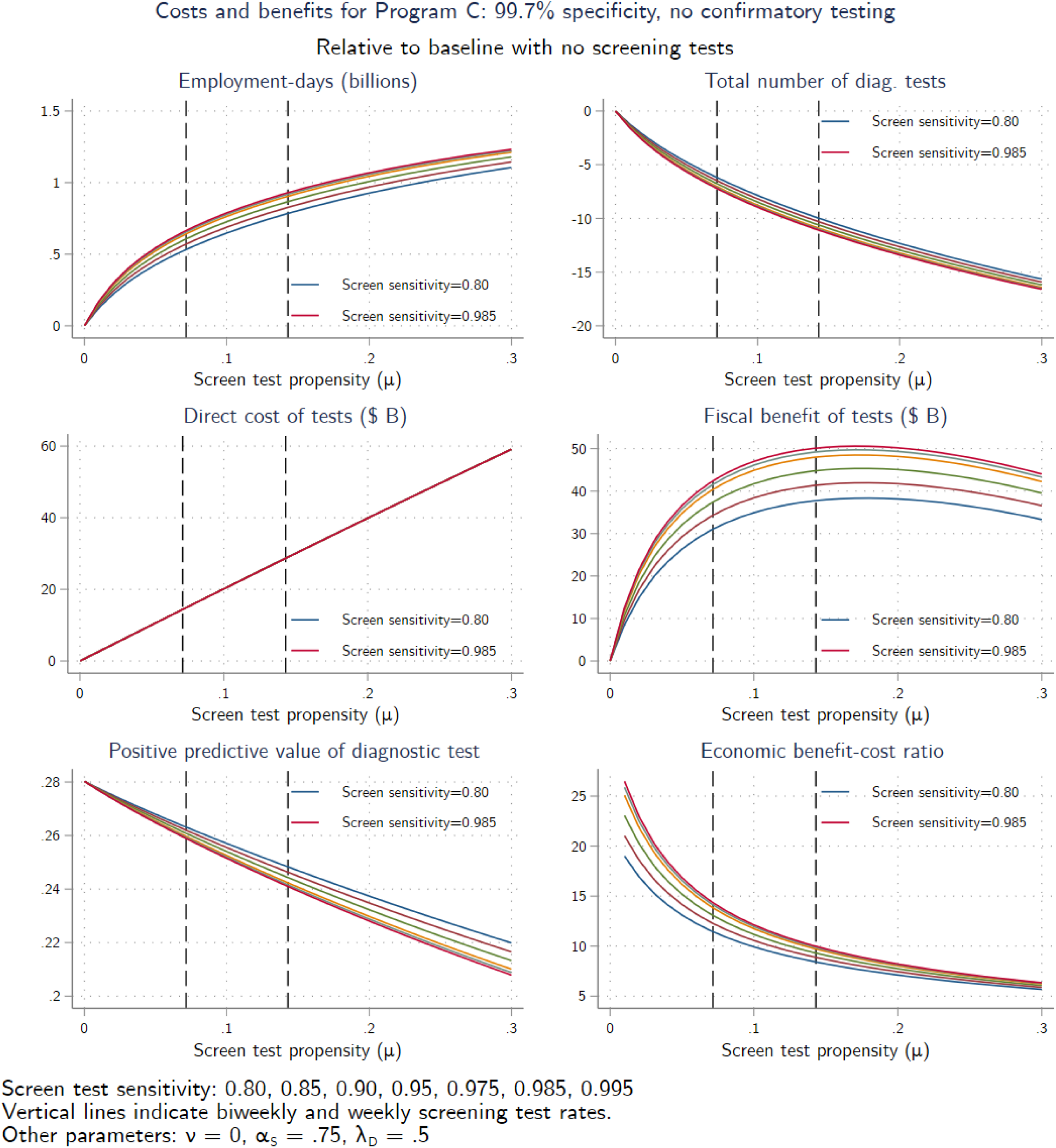
Costs and benefits for program C (no confirmatory PCR testing)

### 3.4 Sensitivity checks

Table 3 and Table 4 summarizes the results of various sensitivity checks.

**Table 4.** Economic and mortality impacts: Arnon et al. (2020) feedback rule

| Testing frequency<br>(days) | Additional testing<br>costs (\$B) | Additional<br>GDP (\$B) | Additional federal<br>receipts (\$B) | Deaths averted<br>(thou) |
| --- | --- | --- | --- | --- |
| *H. Program A* |  |  |  |  |
| 30 | 12 | 23 | 6 | 15 |
| 14 | 26 | 60 | 17 | 35 |
| 7 | 50 | 111 | 31 | 63 |
| 4 | 88 | 152 | 43 | 90 |
| *I. Program B* |  |  |  |  |
| 30 | 13 | 160 | 45 | 65 |
| 14 | 27 | 267 | 74 | 105 |
| 7 | 55 | 354 | 100 | 138 |
| 4 | 97 | 397 | 112 | 154 |
| *J. Program C* |  |  |  |  |
| 30 | 7 | 111 | 32 | 47 |
| 14 | 14 | 179 | 50 | 76 |
| 7 | 28 | 239 | 67 | 105 |
| 4 | 50 | 261 | 73 | 122 |

*Notes:* See the notes to Table 1.

#### Single-stage screening test with 98.5% specificity with no confirmatory testing

A common critique of widespread screening is that low specificity can lead to many healthy individuals, including health workers, needlessly entering isolation (e.g., Pettengill and McAdam (2020)). Panel D in Table 3 considers this case for the 98.5% specificity screening test.

Eliminating confirmatory PCR testing entirely from program A increases the number of healthy people, including healthy workers, in isolation. It also reduces overall adherence because the low-PPV screening test has no follow-up diagnostic testing. These two effects substantially reduce the gains from the screening testing program. In fact, without any confirmatory PCR testing the screening testing program does not pay for itself in most of the cases considered. With no confirmatory testing, there are approximately 1.8 billion proscribed isolation-days with weekly testing, approximately 2.5% of all person-days.

#### Single-stage screening test with 97% specificity with partial confirmatory testing

Panel E modifies program A by considering a screening test with twice the false positive rate of the test in program A. For comparison purposes we hold adherence constant although plausibly it would be lower for the panel E test. The reduced specificity increases the number of healthy individuals proscribed to isolate. Testing costs increase because there are more screening false positives that need confirmation, and isolating so many healthy workers provides an additional drag on GDP. As a result, net economic benefits are less than for program A.

#### Single-stage screening test with 98.5% specificity, partial confirmatory testing, and increased adherence

This scenario, shown in Panel F, modifies program A by increasing adherence from 25% to 50% for those receiving a positive screening test but not taking a confirmatory test. Higher adherence substantially increases deaths averted, GDP, and revenues, and slightly decreases total testing costs because the greater suppression of the virus reduces symptomatic testing costs. Net economic benefits are large, even for biweekly testing.

#### More expensive diagnostic tests

Panel G in Table 3 considers program B (98.5% specificity, universal confirmatory testing) except with a more expensive confirmatory test. The cost of the diagnostic testing is borne by the Federal government so in the model does not affect private decisions and thus does not affect mortality, employment, or GDP. Despite the doubling in the cost of the PCR test, the overall increase in testing cost reduction is modest, for example rising from $56 million for weekly testing in program A (Table 1) to $63 million. The reason is that, with universal PCR confirmatory testing, the expected cost of administering the combined test to an uninfected individual increases only slightly from $5 + .015×$50 = $5.75 for a $50 confirmatory test to $6.50 for a $100 confirmatory test.

#### Alternative economic feedback rule

The results so far use the economic feedback rule in BFMS (2020) (see Appendix 1). As a sensitivity check, we also consider an alternative economic feedback rule taken from Arnon, Ricco, and Smetters (2020). In their rule, activity and employment depend negatively on current cases. Using daily data on activity and caseloads, they estimate an elasticity of employment with respect to cases of -0.0048. The BFMS rule, specialized to depend only on the weekly death rate (not on the slope of deaths or on the unemployment rate), implies a time- and state-dependent elasticity of labor hours with respect to the current death rate. We calculated the BFMS feedback parameter such that the implied elasticity of employment with respect to cases, averaged over the simulation period, equals the Arnon et al. estimated elasticity.^6^

The results are summarized in Table 4, for the three programs in Table 1. The Arnon et al feedback rule is less responsive to the virus than the BFMS rule and induces different dynamics. The estimated gains from the testing programs, both in terms of deaths averted and in terms of GDP gains, are less using the Arnon et al rule than the BFMS rule. All programs, however, continue to yield large net economic and net total benefits. Screening programs B and C are estimated to generate federal receipts exceeding testing costs under the Arnon et al rule, however screening program A does not.

### 3.5. Dynamics for selected scenarios

Figure 9 displays the simulated time path of deaths and quarterly GDP, along with standard error bands and actual deaths, for four counterfactual scenarios. Parts (a), (b), and (c) show respectively programs A, B, and C for a weekly testing rate, and panel (d) shows program C for a four-day testing rate, all computed under the assumptions of Table 1. All cases in Figure 9 have a lower path for deaths and higher path for GDP than the no-screening baseline in Figure 2. Program A slows the spread of the virus but does not suppress it. The other panels, however, approach suppression and two-step testing (program C) at a 4-day cadence essentially suppresses the virus, supporting a strong economic recovery. At a weekly testing cadence, programs B and C could have avoided the second wave of the summer and fall.

**Figure 9.**
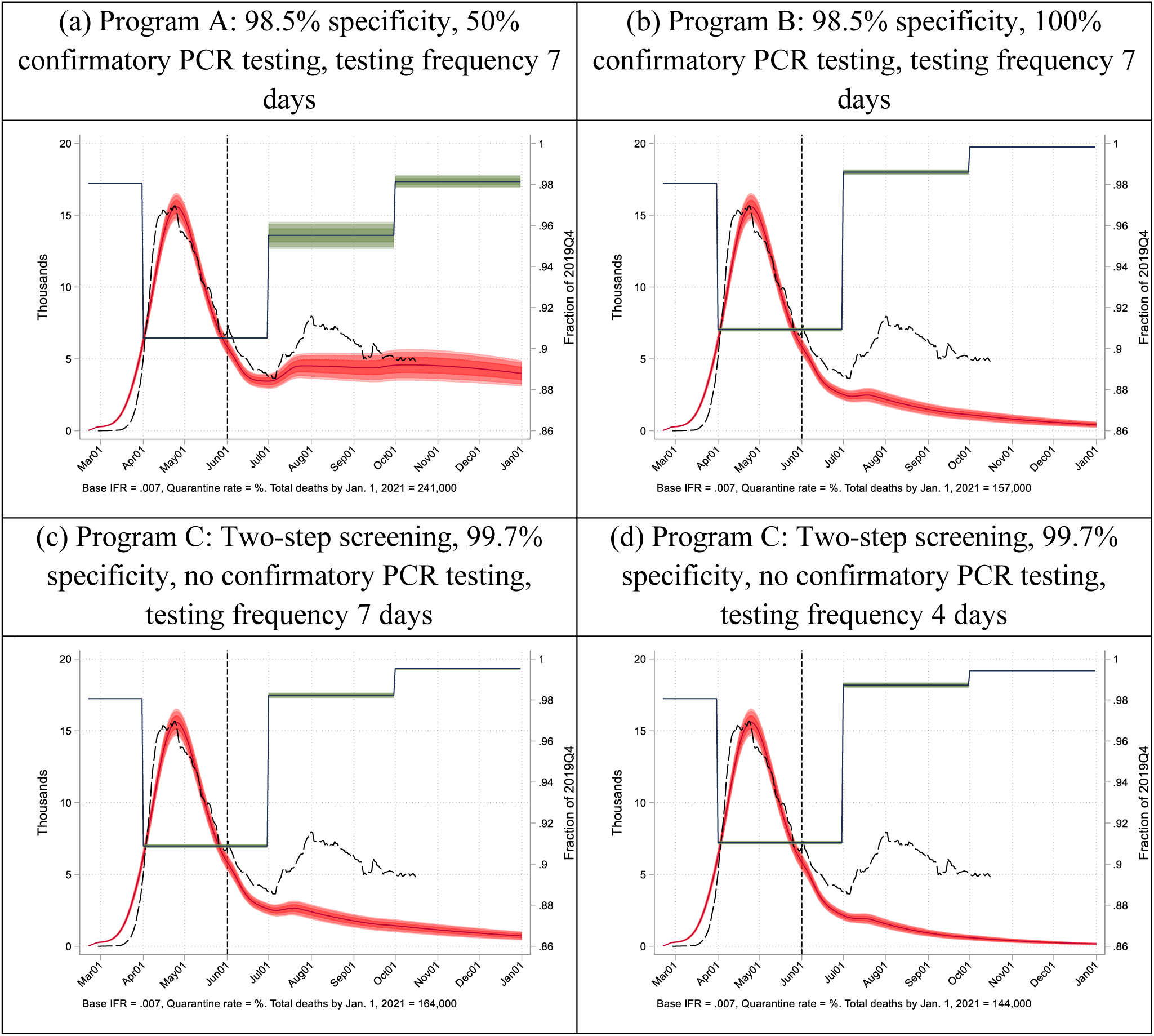
Actual and simulated paths for deaths and GDP. *Notes*: Screening programs A, B, and C are the same as in in Table 1. See the notes to Figure 2.

## 4. Age-Targeted Screening

It might be more efficient to target screening testing based on individual characteristics than having population-wide random screening. Because contacts and mortality differ by age, this section considers screening that is random within an age category with testing rates differing across categories. Specifically, we calculate the age-based testing rates that maximizes net total benefits (economic plus monetized mortality) of the screening test, subject to the constraint that the population-wide screening testing rate equals a given value.

The results of the first calculation – optimized age-specific testing rates – for screening program B (screening test with 98.5% specificity and universal confirmatory PCR testing) are shown in Figure 10, where each line is the probability of testing for a given age. The optimal age-varying testing rates are highest for young adults (ages 20-44) followed by ages 45-64, followed by ages 65-74. These results indicate that the screening testing and isolation is being used to break the chain of transmission from middle-aged adults to the elderly, either through family or service workers serving the elderly. The mortality benefits of this targeting outweigh the economic costs of isolating relatively higher fractions of the working-age population than other ages.

**Figure 10.**
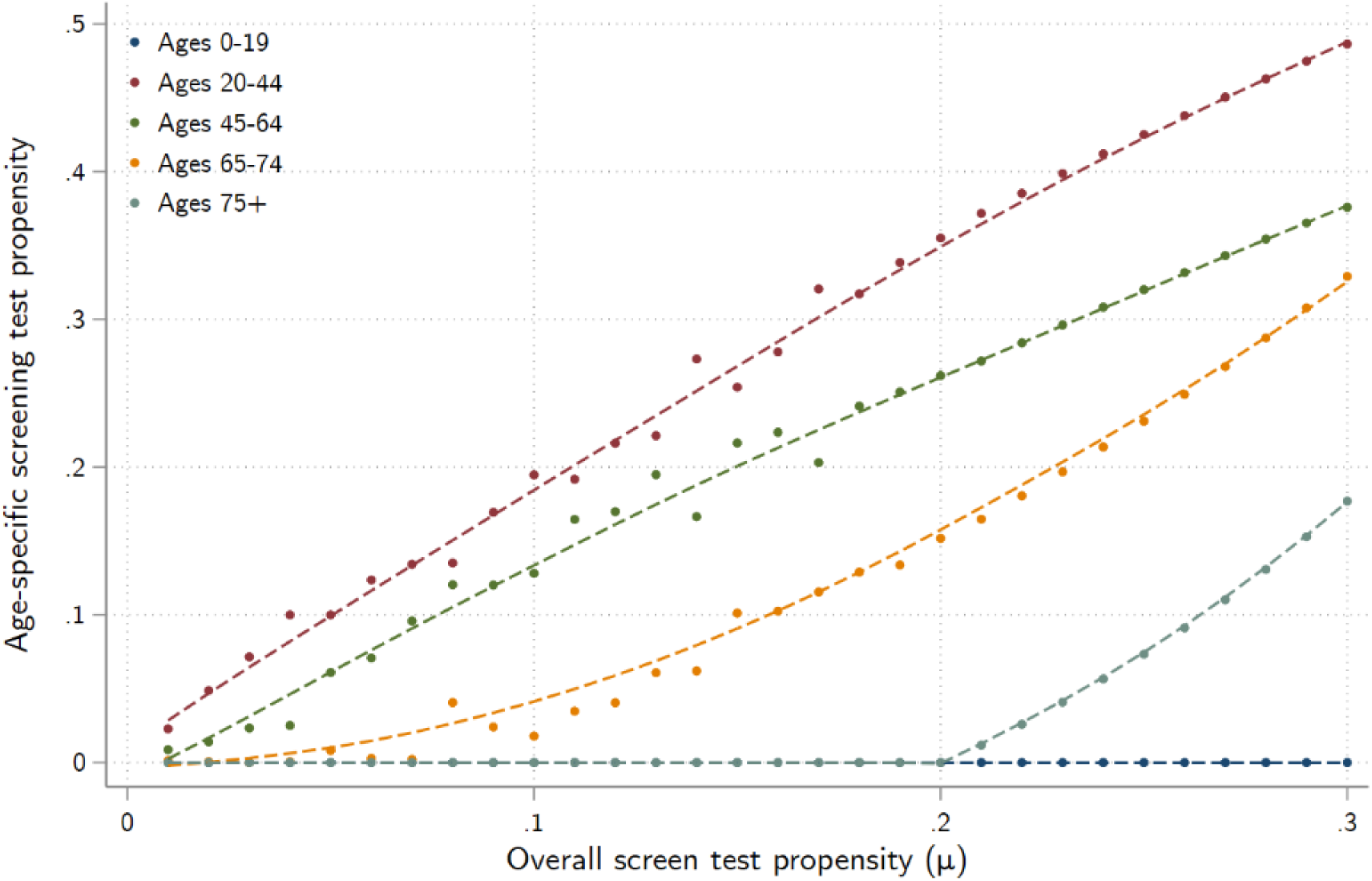
Age-specific screening testing rates that maximize net economic benefits. *Note*: dots are optimization estimates for given overall population testing rate *μ*, lines are smoothed through the estimate by age group.

Figure 11 shows the total net benefits for age-targeted screening testing and, for comparison, for random population screening testing. For small testing rates, there are substantial gains from targeting testing using the unconstrained allocations in Figure 10. Those gains diminish at higher testing rates as the virus is suppressed, however net benefits are always higher with the age-targeted strategy. We note, however, that the costs here do not include developmental and educational costs of children missing school, and including such costs could provide an additional reason to test the young and thus allowing schools to reopen and stay open.

**Figure 11.**
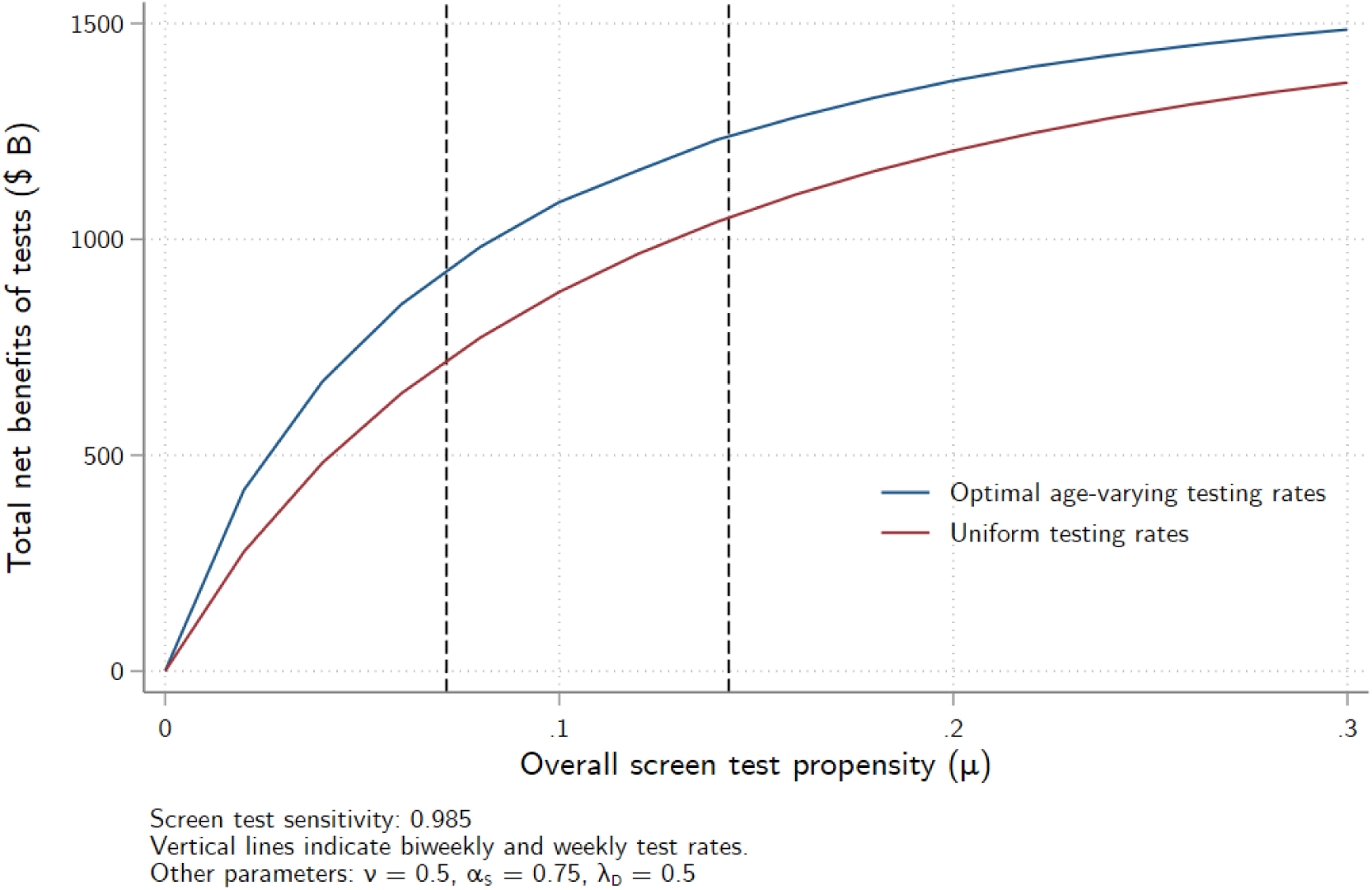
Total net benefits from age-specific and age-blind screening testing.

## 5. Discussion

The specific numerical values reported here depend on the estimated or calibrated epidemiological, economic, and program design parameters, all of which are subject to uncertainty. This uncertainty is reflected in the sensitivity analysis in Section 3.4. For this reason, we stress not any specific set of numerical values, but the robust overall directional conclusion that a screening testing program with high multi-step specificity, designed to evoke high adherence, has the potential both to avert deaths and to provide large economic gains.

There are six main arguments against widespread screening testing (e.g., Pettengill and McAdam (2020)). First, low specificity undercuts the program validity and leads to low adherence with the proscription to isolate if positive. Second, low specificity unnecessarily pulls many healthy workers out of the workforce. Third, because antigen tests have lower sensitivity than PCR tests, many infected individuals would slip through the cracks and undercut the effectiveness of the program. Fourth, if paid for federally, their expense would be massive at a time that the federal deficit is already at a postwar high. Fifth, to be effective they would need to be done at an infeasible scale, such as daily or every other day. Sixth, having a screening program could change behavior, in particularly making individuals who test negative less cautious, for example reducing their willingness to wear a mask.

Our analysis addresses the first five of these concerns. Our results underscore the importance of these first two concerns: in our analysis, the most important parameter is screening test specificity. A screening testing program must have high specificity to be credible and to evoke high adherence. This high specificity can be achieved by two-step testing if the tests are sufficiently independent. The additional costs of two-step testing, even if the second test is a PCR test, are small compared to the benefits, and screening testing with universal PCR confirmatory testing generates large net benefits. Test specificity is typically estimated in a laboratory using a small number of samples, so test specificity in the field could differ substantially from laboratory estimates. Because low specificity undercuts the testing program, this uncertainty underscores the importance of confirmatory testing to increase specificity.

The third concern, sensitivity, is legitimate in theory, but our modeling (like Larremore et al (2020)) finds that even large drops in sensitivity, say to 90%, have a small effect on the epidemiological and economic dynamics. The fourth concern, fiscal sustainability, also is legitimate in theory, but our estimates suggest that the economic gains from suppressing the virus are so large that the testing pays for itself through increased revenue. Regarding the fifth concern, scale, we find that weekly testing in a regime with high compliance comes close to suppressing the virus, and moving to a four-day cadence is highly effective. Weekly testing with a 98.5% specific screening test and universal confirmatory PCR testing would require increasing the number of PCR tests by roughly three-quarters of what they are today; a four-day testing would require more than doubling PCR testing capacity.

Our analysis does not tackle the final concern, that testing could induce more risky behavior. With that caveat, it is not self-evident that this must be the case. Individuals undertake social distancing and masking to self-protect, to protect others, and to conform to local norms and laws. Testing negative in the morning does not reduce the incentive to self-protect during the day. The effect on behavior of testing positive is ambiguous: altruism would lead one to reduce contacts even if not isolating, but no longer worrying about one’s own health while caring little about the health of others could increase risky behavior. Empirical research on this effect is needed.

The analysis here relies on several simplifications and thus has multiple caveats. On the economic side, the GDP concept in this paper is private sector output and it does not include any of the deficit-financed emergency support measures passed during the pandemic. The model is estimated for the entire United States, so it necessarily misses regional heterogeneity, nor does it model frictions or adjustment dynamics other than those arising from epidemiological dynamics.

On the epidemiological side, we do not incorporate contact tracing because of the general lack of timely contact tracing currently in the United States. Contact tracing would enhance the benefits of increased testing, especially from additional PCR tests within the public health system, because more cases would be identified; however, it would also increase costs, and a next step is to undertake a cost-benefit assessment of combining screening testing with enhanced contact tracing (also with partial adherence). Another is that the model does not differentiate between high- and low-risk workplaces. Just as we found benefits to age-based targeting of screening, it is plausible that there would be benefits to targeting screening based on the amount of work-based contacts. In addition, we do not model pooled testing. If antigen tests are in short supply, they potentially could be pooled, for example at the household level, with confirmatory antigen and/or PCR testing. This too has the potential for increasing the benefit-cost ratios. Nor do we model incentives to increase adherence to instructions to isolate, although our sensitivity analysis confirms that increased adherence serves further to suppress the virus. For some, adherence is a behavioral response, but for others, adherence is difficult because of living circumstances (room-mates, extended family sharing housing) or financially because of missed work, and in those cases policy interventions might help those who wish to adhere to do so. Further, we do not consider repeat PCR testing because of existing constraints on PCR testing capacity, however repeat PCR testing might be useful for identifying the stage of infection thereby fine-tuning personalized isolation times and reducing the aggregate burden of isolation (Kissler et al (2020)).

Finally, our study of the economic benefits of COVID-19 screening tests does not consider the public health benefits of the data generated from such a testing program for disease surveillance purposes. (We note that testing for both diagnostic and public health surveillance purposes is already routinely employed for both seasonal influenza and detection of novel strains of influenza A.) Although at-home screening test results would not get into a public data system, universal confirmatory PCR testing would increase data coverage by overcoming the current selection into diagnostic testing of the symptomatic. Thus, the testing regimes considered here would allow for much more timely and fine-grained analysis of the response of COVID-19 prevalence and transmission to a wide range of public health interventions and disease mitigation strategies than is possible with current diagnostic testing data. Presumably, consideration of the utility of the data generated from a widespread screening testing regime in shaping the design of effective and low-cost mitigation measures would add to the economic benefits.

## Data Availability

All data used ihis paper is derived from publicly available sources and available from the authors upon request.

## Appendix 1

This appendix provides more details on the model, which extends the model developed in Baqaee, Farhi, Mina, and Stock (2020). Our model departs from BFMS in several important respects. First, we extend the model to include both a screening test regime and a diagnostic test regime. Second, we assume that individuals are instructed to isolate upon receiving a terminal positive test, either a screening test with no confirmatory test or a positive diagnostic test. Individuals awaiting diagnostic test results are instructed to quarantine. Third, we distinguish between individuals who have recently recovered and fully recovered from the disease to capture that individuals may still test positive on a PCR test after they are no longer infectious. Finally, we allow for imperfect adherence to quarantine and isolation.

### The Epidemiological Model

There are five age groups indexed by *a*, representing ages 0-19, 20-44, 45-64, 65-74, and 75+. There are 66 private sectors in the economy indexed by *i*. Individuals are either *S* (susceptible), *E* (exposed), *I* (infected), *R* (recently recovered), *F* (fully recovered), or *D* (dead). In addition, individuals who are not dead are either actively circulating (*A*), awaiting diagnostic test results (*D*), awaiting screening test results (*S*), or in isolation following a positive test (Q). Thus, the population is partitioned into 21 states. For example, *SA*_2_ *SS*_2_, *SAD*_2_, and *SQ*_2_ denote the number of persons aged 20-44 that are susceptible and actively circulating, susceptible and awaiting screening test results, susceptible and awaiting diagnostic test results, and susceptible and in isolation, respectively. We assume that the recovered (either recently recovered or fully recovered) are immune through the end of our simulation period.

The rates of screening and diagnostic testing are given by the parameters *μ, ρ*_0_, and *ρ*_1_, described in Table 2. We assume that these parameters are equal to zero in the estimation period of our model, which runs through June 1^st^, and thereafter calibrated according to the main text of this paper.

The state variables (i.e. SA) are all five-dimensional vectors. Let *X*_*a*_ denote the *a*^th^ element of any state *X* (the *a*^th^ age group). The epidemiological side of the model has 21 transition equations:

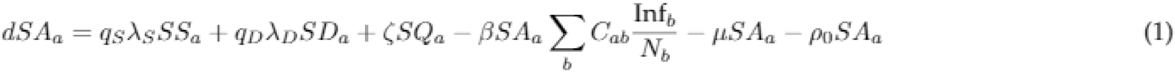

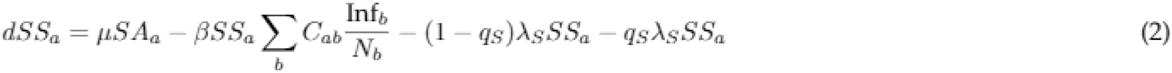

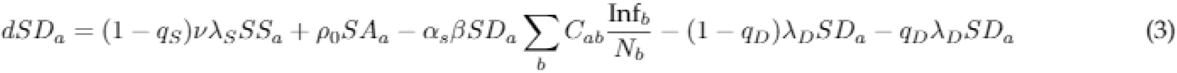

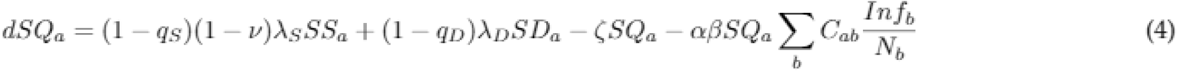

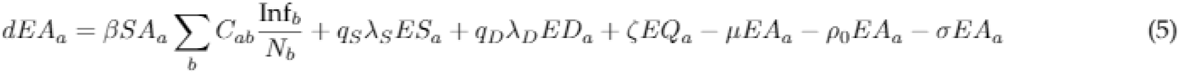

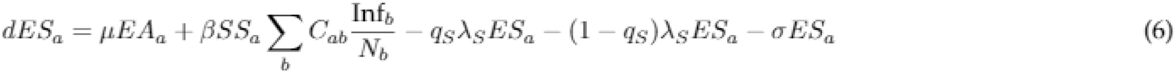

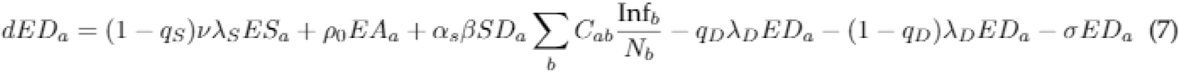

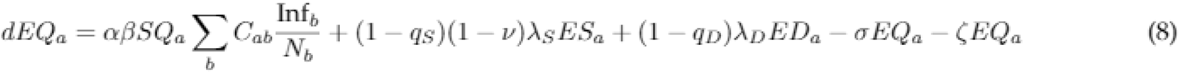

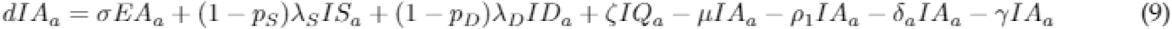

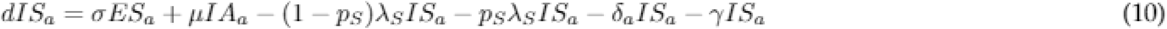

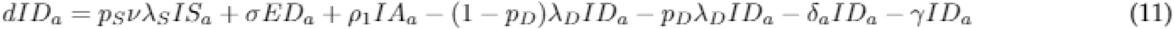

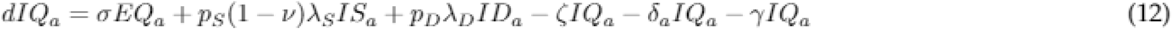

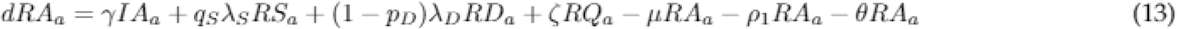

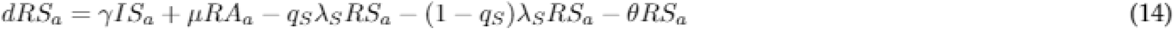

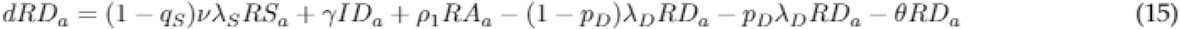

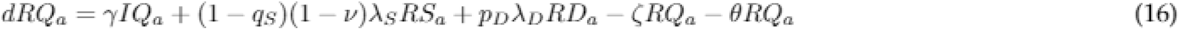

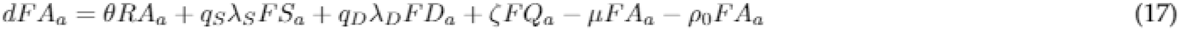

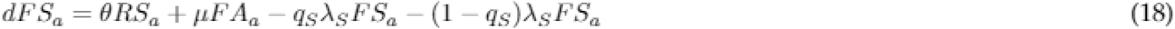

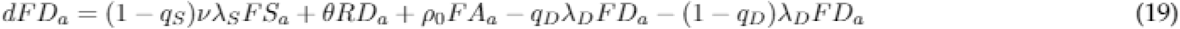

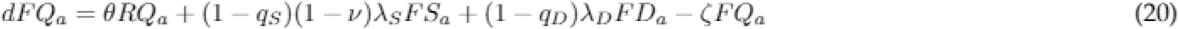

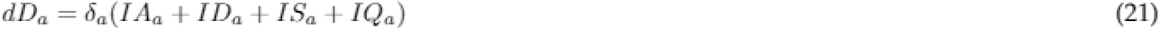

where *N*_*a*_ denotes the number of individuals of age a (summing across all 21 states) and *Inf*_*a*_ denotes the effective number of infected individuals actively circulating, *Inf*_*a*_ = *IA*_*a*_ + *IS*_*a*_ +*α*_*S*_*ID*_*a*_ + *αIQ*_*a*_. In this final expression we treat the signal value of taking a diagnostic test as being the same as receiving a positive screening test (these would be the same for the screening-test positives taking a confirmatory PCR test), so non-adherence with quarantine is the same as non-adherence with a terminal positive screening test.

The parameter *α* is a weighted average of the parameters *α*_*S*_ and *α*_*D*_, which are the isolation adherence rates for those who received screening tests and diagnostic tests, respectively. The weights are endogenously determined and given by the relative share of those instructed to isolate who arrived from screening tests versus diagnostic tests. Thus, *α* is the effective adherence rate of those in isolation. If those in isolation mainly through the screening test regime, then *α* will be close to *α*_*S*_, the quarantine adherence rate for the screened population.

Given the parameters appearing in equations (1) through (21) above and a set of initial conditions, the model is straightforward to solve in discrete time by forward iteration. The unit of time is a single day and the model is solved 12 steps per day.

#### The Contact Matrix

The contact matrix *C* describes the expected number of contacts between each age group in the population. An actively circulating individual of age *a* who interacts with an individual of age *b* has an instantaneous infection probability of *β* times the probability that the age-*b* individual is infected. The probability that an individual of age a is infected in a given period is therefore given by summing across all their contacts. We distinguish between contacts that are made at home, at work, and elsewhere. The contact matrix is time-varying, and can change due to, for instance, NPIs put in place by the government or personal behavioral adaptations to avoid contracting the virus. We have:

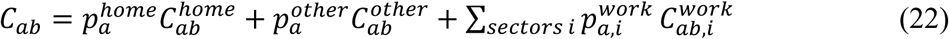

Where 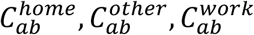 indicate the expected number of contacts in each of home, work, and other environments, conditional on being at home, at work, or elsewhere. The parameters 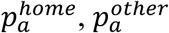 and 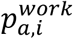 indicate the probability that an age-a individual is at home, at work, or elsewhere. We note that 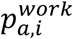 is the fraction of employed in the indicated sector:

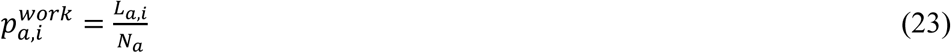

where *L*_*a,i*_ is the number of workers of age *a* employed in sector *i*.

See Sections 1.2 and 2 of BFMS (2020) for more information on the construction and historical estimation of the contact matrix.

#### Behavioral Feedback and Control Rule

The behavioral component of this model endogenously determines the contact matrix in our simulation period (i.e. after June 1). This portion of the model is unchanged from BFMS 2020. For completeness, we will briefly describe the key elements of this control rule here.

In our simulation period (June 1^st^ through December 31^st^), we assume that the contact matrix responds endogenously to changes in the course of the pandemic. We implement this using a linear proportional-integral-derivative (PID) control rule, in which feedback depends on current deaths, the 14-day change in deaths, the current unemployment rate and the integral of the unemployment rate. The linear PID control rule can be expressed as:

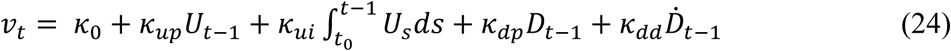

where *U*_*t*_ is the unemployment rate and 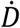 is the time derivative of the death rate. Both *U*_*t*_ and 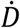 are generally unknown, available only with time aggregation and/or with reporting lags. We therefore use the 14-day average of the unemployment rate, the cumulative daily unemployment rate since March 7^th^, deaths over the previous two days, and the 14-day change in the two-day death rate for the various terms on the right-hand side of this equation.

The PID controller determines a sequence of sectoral labor supply shocks, shifted by the GDP-to-risk index:

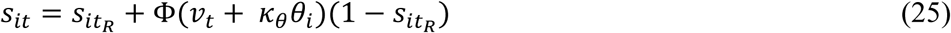

where *s*_*it*_ is labor hours *Lit* in sector *i* at date *t* as a fraction of labor hours prior to the pandemic (i.e. February 2020), *t*_*R*_ is the date of the beginning of the simulation period (June 1^st^), and Φ is the cumulative Gaussian distribution (which plays no role except as a sigmoid to constrain the controller between 0 and 1).

The term *θ*_*i*_ is the GDP-to-risk index:

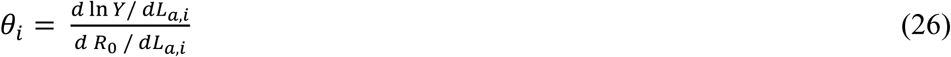

The GDP-to-risk index can be interpreted as measuring the ratio of the marginal contribution to output, relative to the marginal contribution of *R*_0_, from an additional worker of age *a* returning to work in sector *i*. Up to scale, the GDP to risk index does not depend on epidemiological parameters except the contact matrix. The units of *θ* are not meaningful, so we standardize it to mean zero and unit variance across sectors (equally weighted).

Thus, the controller effectively alters the work contacts component of the contact matrix. Similarly, we can think of the controller as generating a sequence of labor supply shocks that can be used to back out GDP using Hulten’s theorem as a first-order approximation:

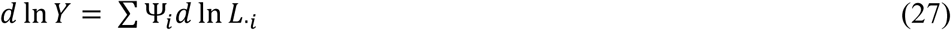

Where the subscript · denotes summation over ages and Ψ_*i*_ denotes the labor income share for sector *i*.

## Acknowledgements

We thank Alan Auerbach, Daniel Larremore, Larry Summers, and seminar participants at Johns Hopkins University, the Federal Reserve Bank of Boston, and the Harvard Kennedy School for helpful discussions and/or comments. Droste and Stock acknowledge research support under NSF RAPID Grant SES-2032493.

## Footnotes

1 See Arnon et al (2020), Goolsbee and Syverson (2020), Chetty et al (2020), and Gupta, Simon, and Wing (2020) and the literature cited in those papers.

2 The positive predictive value is the probability of being infected conditional on testing positive. By Bayes Law, the PPV depends on the specificity and sensitivity of the test and on the population rate of infection.

3 These costs and accuracy rates are those of the Abbot Laboratories BinaxNOW™antigen test (FDA (2020)). Additional estimates of test performance and costs are available in Table 2 of Silcox et. al. (2020).

4 Meta-analyses of influenza antigen tests estimate specificity of 98.2% (Chartrand et al (2012)) and 98.4% (Antoniol et al (2018)), see Pettengill and McAdam (2020)). These specificities are close to the BinaxNOW™ specificity of 98.5%. In this light, the assumed 80% specificity for the first stage is conservative.

5 An early focus of this literature concerned the macroeconomic and epidemiological effects of lockdown and re-opening policies. Eichenbaum, Rebelo, Trabant (2020a) augment a standard New Keynesian macroeconomic model with a SIR-type model of disease transmission, characterize the relationship between consumption/labor supply decisions and disease transmission, and study the effects of simple lockdown policies. Acemoglu, Chernozhukov, Werning, and Whinston (2020) study a multi-group SIR model where infection, hospitalization, and fatality rates vary between groups and characterize optimal age-varying lockdown policies. This literature has expanded to include other non-pharmaceutical interventions, see Baqaee, Farhi, Mina, and Stock (2020a). See BFMS (2020b) for additional references.

6 Specifically, to align the BFMS rule with Arnon et al, we set *κ*_*up*_, *κ*_*ui*_, and *κ*_*dd*_ in Appendix equation (24) to 0 and solved for the value of *κ*_*dp*_ for which the average model-implied elasticity matched the Arnon et al elasticity of employment with respect to cases. That elasticity is related to the elasticity of labor hours with respect to deaths, computed from Appendix equations (24) and (25), as, 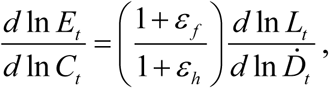 where *E*_*t*_ is employment, *L*_*t*_ is labor hours, *C*_*t*_ is cases,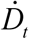 is daily deaths, *εf* is the elasticity of the case fatality rate with respect to cases, and *εh* is the elasticity of weekly hours with respect to employment. The elasticity *εh* was estimated from aggregate US data on hours and unemployment, and the elasticity *εf* was estimated from data on the cases and deaths from July 1 – October 22, 2020 (a period in which the number of tests were roughly constant).

